# Machine Learning-Based Models for Imputing Missing Values of Key Predictors of Important Clinical Outcomes in a Large-Scale Registry of Acute Myocardial Infarction

**DOI:** 10.1101/2025.05.29.25328603

**Authors:** Haoyun Hong, Remy Poudel, Asishana Osho, Kathie Thomas, Shen Li, Abhinav Goyal, Jennifer L. Hall, Juan Zhao

## Abstract

**Background:** Missing data is a frequent limitation of database research. Application of machine learning-based models is a promising approach to overcome this limitation. This study aims to evaluate methods for imputing three critical but sometimes missing predictors of important clinical outcomes (e.g. mortality) in hospitalized patients presenting with acute myocardial infarction (AMI): cardiac arrest prior to arrival, heart failure on first medical contact (FMC), and cardiogenic shock on FMC.

**Methods:** The study included 219,146 individuals participating in the American Heart Association Get With The Guidelines® Coronary Artery Disease (GWTG-CAD) Registry from 2020 to 2022. We trained machine learning models along with logistic regression model to predict cardiac arrest prior to arrival, heart failure on FMC and cardiogenic shock on FMC through a nested cross-validation with a calibration layer. Important features are identified through Shapley additive explanations (SHAP) and model performance are evaluated on different demographic subgroups.

**Results:** The mean (SD) age was 65 (18) years, with 32.7% Female; 71.2% individuals were White, and 13.5% individuals were Black. For cardiac arrest, XGBoost and LightGBM achieved AUROC scores of 0.920 (95% CI: 0.915 – 0.924) and 0.922 (95% CI: 0.918 – 0.927). For cardiogenic shock, XGBoost and LightGBM achieved AUROC scores of 0.912 (95% CI, 0.907 - 0.918) and 0.913 (95% CI, 0.908 - 0.918). For heart failure, XGBoost and LightGBM achieved AUROC scores of 0.824 (95% CI, 0.818 - 0.829) and 0.826 (95% CI, 0.818 - 0.833). Vital signs, lab values, age, and AMI type are among the most influential features for predicting these clinical presenting variables.

**Conclusions:** We demonstrated machine learning techniques, particularly XGBoost and LightGBM, have proven effective in imputing missing clinical data in cardiovascular studies. The models improve the data power and quality of datasets by accurately imputing major cardiac events, with implications for enhancing clinical and research practices.

## INTRODUCTION

As data and predictive analytics are increasingly used in healthcare, the integrity and completeness of clinical data become crucial to the success of using such tools to advance patient care and clinical research.^1–3^ However, missing data remains a prevalent challenge for large data sources such as clinical registries and electronic health records. The reasons can be attributed to multiple factors, including changes in data collection methods or forms, and the nuanced nature of clinical encounters.^4^ Missing clinical data and fragmented patient records undermine the utility of large datasets in research and quality improvement by reducing statistical power and producing biased outcomes.

The conventional approach to addressing this issue has been limited to removing patients’ records with missing data, or using traditional statistical imputation techniques, ^5–9^ such as single imputation of means, medians or modes, and multivariate imputation by chained equations. However, these methods have been used more frequently for continuous variables such as lab values. Inferring data about dichotomous variables is more challenging, as imputation in this context requires detailed consideration of the clinical context for each unique patient.

The American Heart Association’s Get With The Guidelines® – Coronary Artery Disease (GWTG-CAD) Registry is large AMI registry used for quality improvement and clinical research. Several demographic, clinical, and hospital variables are collected for each patient from the time of first medical contact (FMC) in the prehospital setting with emergency medical services (EMS), as well as in the hospital through discharge, and several in-hospital outcomes (e.g. mortality) are documented. Three patient variables at the time of FMC are particularly strong predictors of in-hospital outcomes, namely cardiac arrest, heart failure, and cardiogenic shock. ^10–12^ Yet, documentation of these three key presenting conditions is sometimes missing in the GWTG-CAD registry. In this study, we leveraged machine learning algorithms to impute missing values of cardiac arrest, heart failure, or cardiogenic shock among patients admitted to the hospital with STEMI or NSTEMI. First, we evaluated the predictive accuracy of machine-learning (ML) algorithms such as XGBoost and LightGBM and compared them with standard imputation methods. Second, we utilized a framework, called Shapley additive explanations (SHAP),^13^ to identify the most influential factors for inferring these three clinical indicators. Third, we evaluated the performance of models on multiple demographic subgroups.

## METHODS

### Data Sources and Collection (Get with the Guidelines Coronary Artery Disease Cohort)

The American Heart Association GWTG – CAD® Registry is a large, international clinical registry aimed at improving the quality of care of AMI patients. Participating hospitals contribute clinical data of consecutive patients admitted with AMI.^14,15^ Detailed information about this registry and data elements collected via the case report form can be found at: https://www.heart.org/en/professional/quality-improvement. Data were used primarily at local sites for quality improvement; therefore, sites were granted a waiver of informed consent under the common rule. The data collection and coordination for GWTG programs are managed by IQVIA. Deidentified data from the AHA GWTG-CAD registry are available through the AHA Precision Medicine Platform. Requests to access the data should be made by contacting AHA as instructed at https://www.heart.org/en/professional/ quality-improvement/quality-research-and-publications/ hospital-level-research.

### Study Period and Population

Data from the AHA GWTG-CAD registry was accessed on June 30, 2023. The study population included all patients with a principal final/discharge diagnosis of STEMI or NSTEMI, who were admitted to one of the GWTG-CAD hospitals from January 1, 2020, to December 31, 2022 (Figure 1). Records with missing sex, race/ethnicity, or discharge status are excluded from the study.

**Figure 1.**
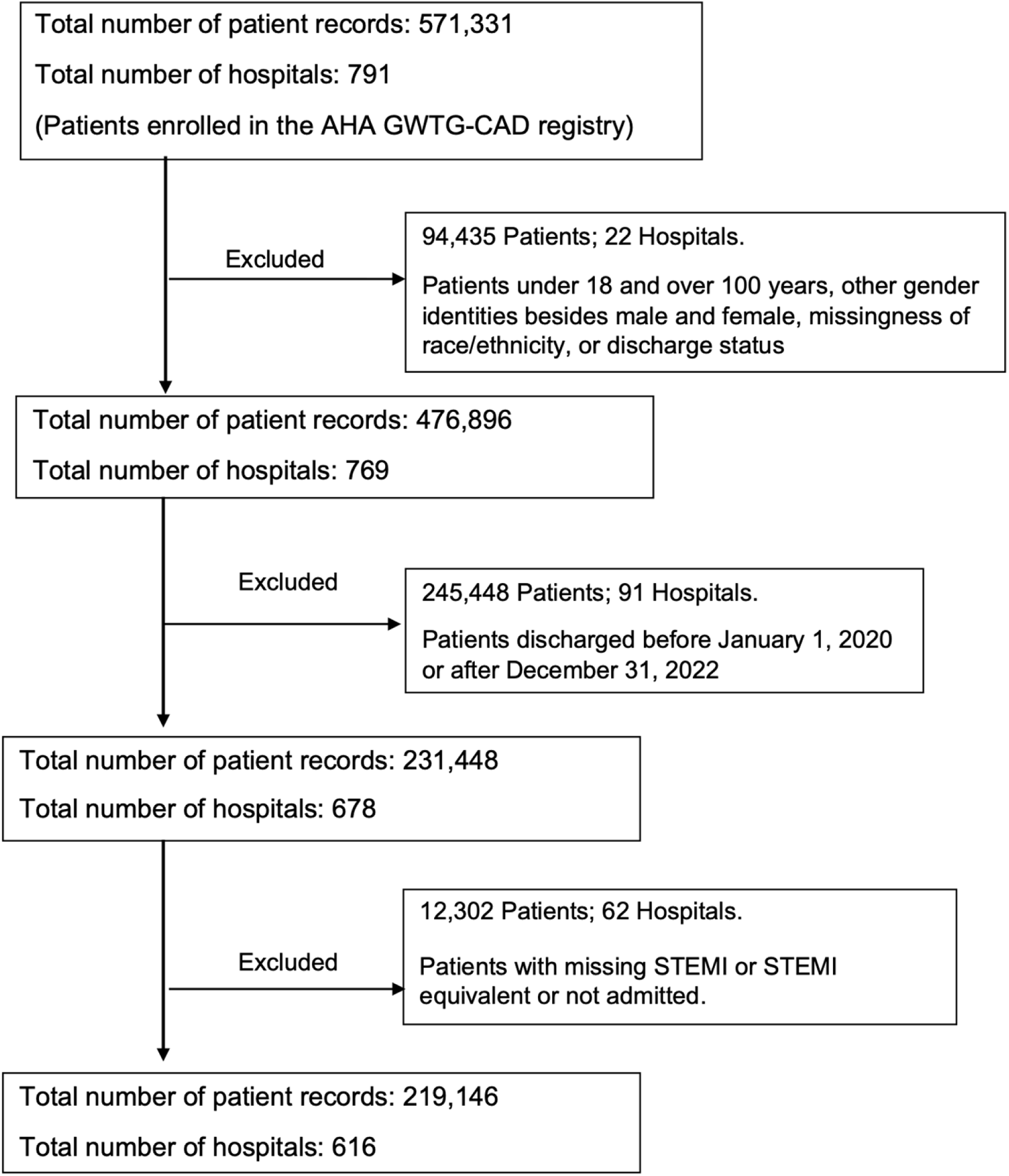
Inclusion and Exclusion Criteria of Study Population.

### Outcomes and Data Variables for prediction

The primary outcomes are presence or absence of three critical clinical conditions: cardiac arrest prior to arrival, heart failure on FMC, and cardiogenic shock on FMC. These outcomes were categorized as a binary variable (entered for each patient as “yes” or “no”). We used a range of predictors (features) including demographics, medical history, vital signs, and lab values upon hospital admission, medications, and procedures during hospitalization. We excluded variables that are less relevant for prediction (e.g., date of admission, hospital characteristics), as well as important downstream outcomes (e.g. mortality) that these variables would be used to predict. Furthermore, we excluded any variables with more than 40% missing rate. For continuous variables, we used median value for imputation. All categorical variables are one-hot-encoded. A complete list of final features included in the ML models is shown in Supplementary Table 1.

### Statistical Analysis

Categorical variables were summarized using percentages. Continuous variables were summarized using medians with interquartile range. Statistical significance was assessed at α < 0.05.

### Machine Learning Models

We applied four machine learning models including (1) logistic regression (used as reference model), (2) random forest, (3) extreme gradient boosting machine (XGBoost), and (4) Light gradient Boosting Machine (LightGBM). XGBoost Trees and LightGBM are two effective implementations of gradient boosting trees (GBT) that have been applied previously to predict health outcomes and have demonstrated strong performance.^16–18^ We trained each model on each of the outcomes and assessed the performance using the metrics listed below.

### Model Training, Calibration, and Evaluation

To evaluate the imputation method, for each of the three outcomes, we obtained a separate, complete dataset. We then adopted a nested stratified cross-validation (CV) process, complemented by a calibration layer and a 5% hold-out test set to ensure robust evaluation (Figure 2). We used the nested CV to ensure that the data for hyperparameter tuning, calibration, and testing were strictly separated. The complete dataset is initially divided using a 5-fold outer CV. For each of the five iterations, a different fold serves as the test set, and the remaining folds form the training set. Within each iteration of the outer CV, there’s an inner 5-fold CV applied to the training set. This inner CV is used for hyperparameter tuning – models are trained and validated across the five folds to identify the best hyperparameters set. To ensure precise calibration of our models, an isotonic regression is applied in the validation set of the calibration layer to ensure the models are well-calibrated. The cut-off threshold was determined in the test set of the calibration layer using F1 scores, which is the harmonic mean of the precision and recall. This calibrated model was then evaluated on the test set of the outer CV fold by the area under the receiver operating characteristics (AUROC), as well as other metrics including accuracy, sensitivity, specificity, positive predictive value, and negative predictive value.

**Figure 2.**
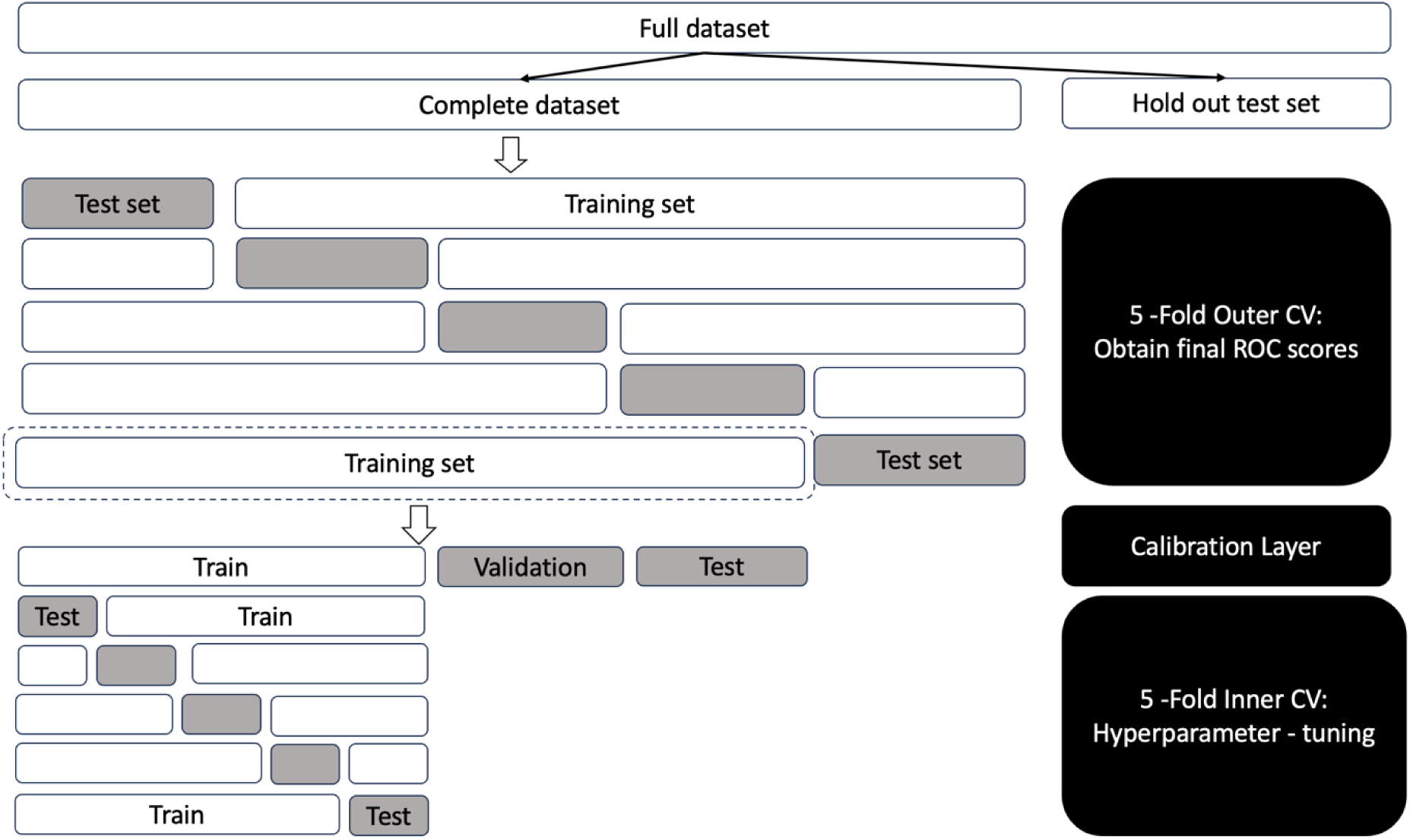
Study Design: For each of the three outcomes, a full dataset is obtained after removing records with missing outcome. Then the dataset is split into complete dataset (95%) and hold out test set (5%). A nested CV with a calibration layer is then conducted on the complete dataset with the inner 5-fold inner CV used for hyperparameter tuning and 5-fold outer CV used for obtaining final ROC scores and other performance metrics after model calibration in the calibration layer where thresholds are determined.

### Feature Importance

To demonstrate the feature importance for complex models such as XGBoost and lightGBM, we used an outstanding approach – Shapley additive explanations (SHAP). The SHAP explanation method computes each feature’s contribution (i.e., Shapley value) to explain the prediction of an instance through a coalitional game theory. We applied SHAP to XGBoost models to first generate SHAP values for all features, summarized the impact of the top 20 features on the model output through beeswarm plots, and analyzed the impact of top 10 features in two specific instances through waterfall plots using the publicly available SHAP API^19^.

### Subgroup analysis

To evaluate any bias that was produced in model prediction, we further assessed the prevalence rate, false negative rate (FNR), and false positive rate (FPR) across different sex and race/ethnicity groups in the 5% hold-out test set. We chose the optimal model that achieved most outstanding performance for this assessment.

### Implementation Details

Machine learning models, cross-validation, and evaluation metrics were implemented using Sci-kit Learn 1.3.2. Data analysis and statistical analysis were performed using Python version 3.11.5 and the open-source software R (v4.2.0), R Foundation for Statistical Computing, Vienna, Austria). SHAP interpretation was implemented with *shap* python package.

## RESULTS

### Patient Baseline Characteristics

A total of 219,146 patients from 616 hospitals participating in the American Heart Association GWTG-CAD Registry were included in the analysis. A summary of demographics and clinical characteristics is shown in Table 1. Among them, 6,994 (3.2%) patients had cardiac arrest prior to arrival, 26,290 (12.0%) patients had heart failure on FMC, and 9,218 (4.2%) patients had cardiogenic shock on FMC. 53.8% of patients are missing cardiac arrest prior to arrival status, 7.0% of patients are missing cardiogenic shock on FMC and 3.2% of patients are missing heart failure on FMC (Supplementary Table 2).

**Table 1.**
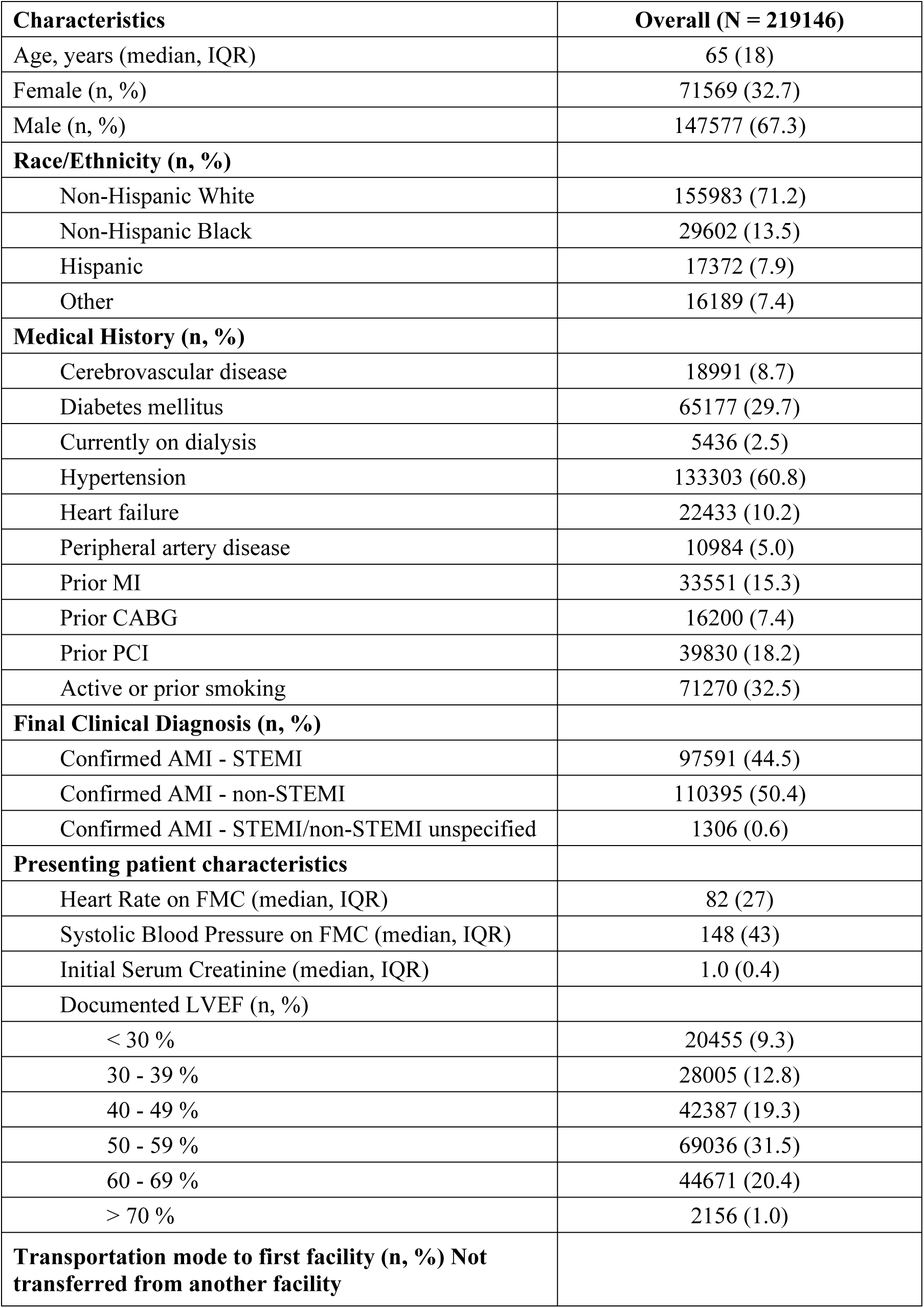
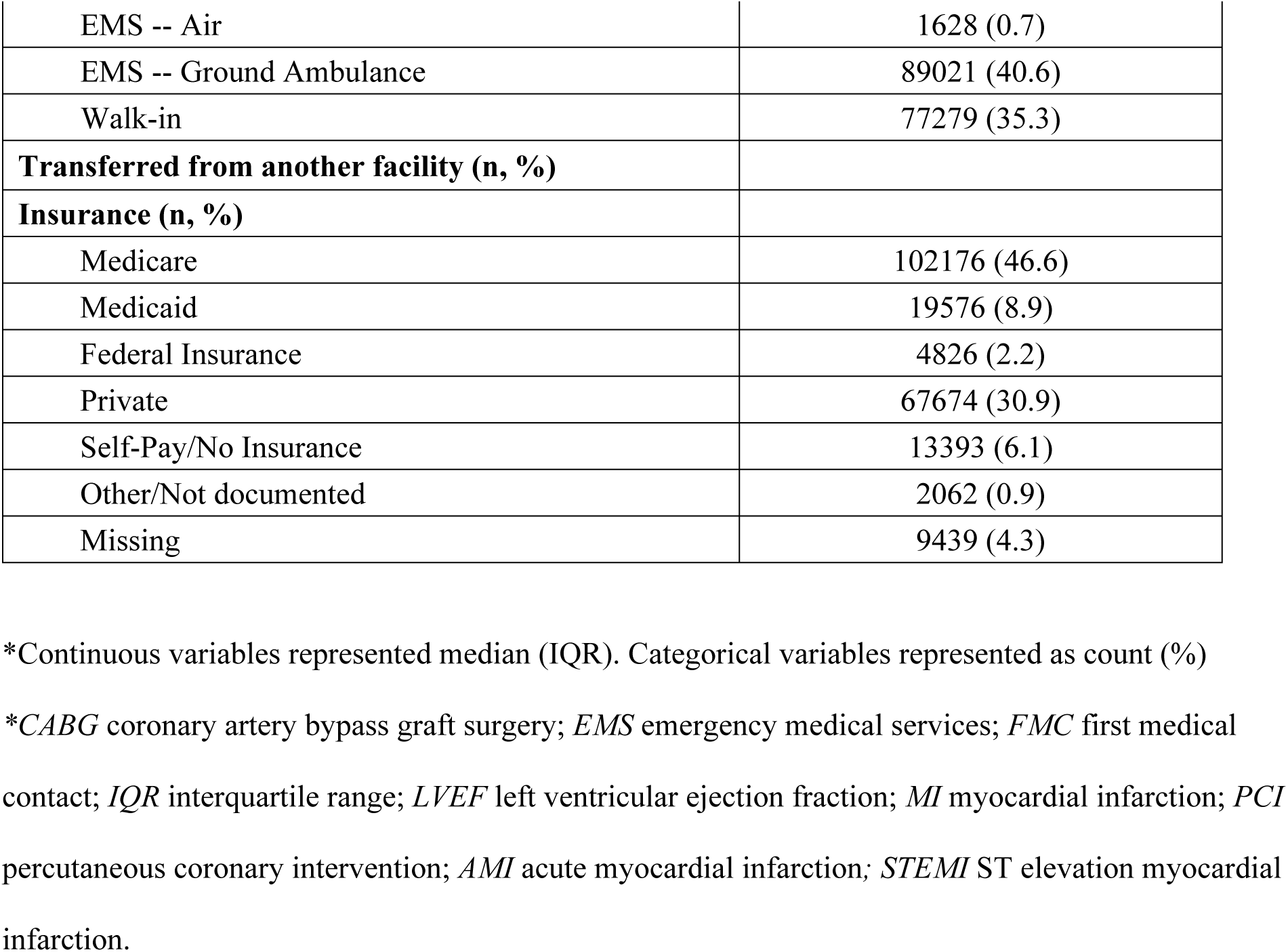
Baseline Characteristics of cohort.

### Model Performance

In performance for imputing three distinct outcomes, XGBoost and LightGBM achieved higher AUROC scores than logistic regression and random forest models. In predicting cardiac arrest before arrival, XGBoost and LightGBM achieved AUROC scores of 0.920 (95% confidence interval [CI], 0.915 – 0.924) and 0.922 (95% CI, 0.918 – 0.927) respectively (Supplementary Table 3). In predicting cardiogenic shock on FMC, XGBoost and LightGBM achieved AUROC scores of 0.912 (95% CI, 0.907 - 0.918) and 0.913 (95% CI, 0.908 - 0.918). Similarly for predicting heart failure on FMC, XGBoost, and LightGBM achieved higher AUROC scores of 0.824 (95% CI, 0.818 - 0.829) and 0.826 (95% CI, 0.818 - 0.833). Notably, XGBoost and LightGBM performed similarly across many key metrics including accuracy, sensitivity, specificity, negative predictive value, and positive predictive value.

### Feature Importance

The XGBoost models were finally used in identifying top 20 features that significantly influenced the prediction of cardiac arrest before arrival, cardiogenic shock, and heart failure at the time of first medical contact (FMC). The influence of these features was ranked according to their contribution to the model’s predictions, based on the average SHAP values (Figure 3a-3c).

**Figure 3.**
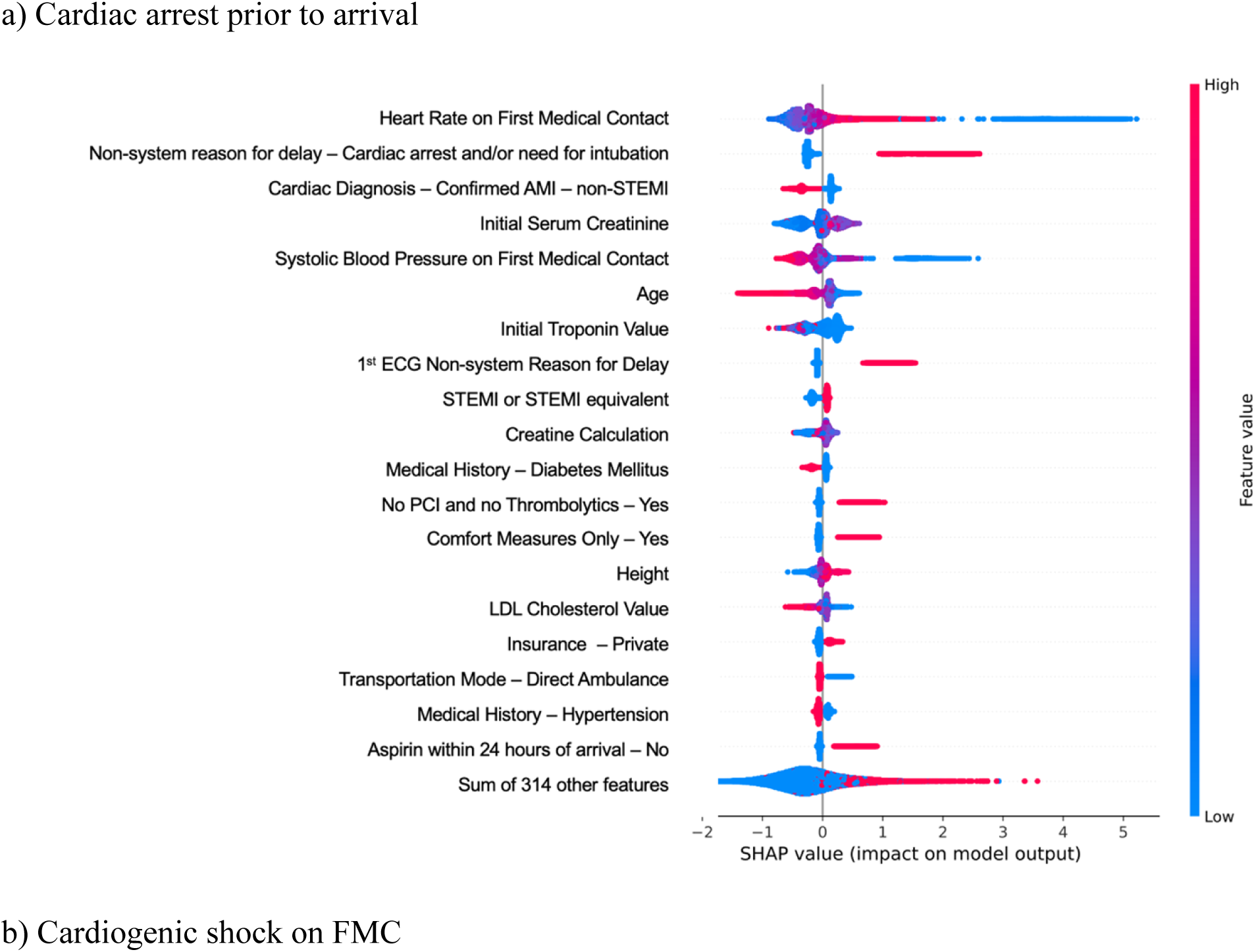
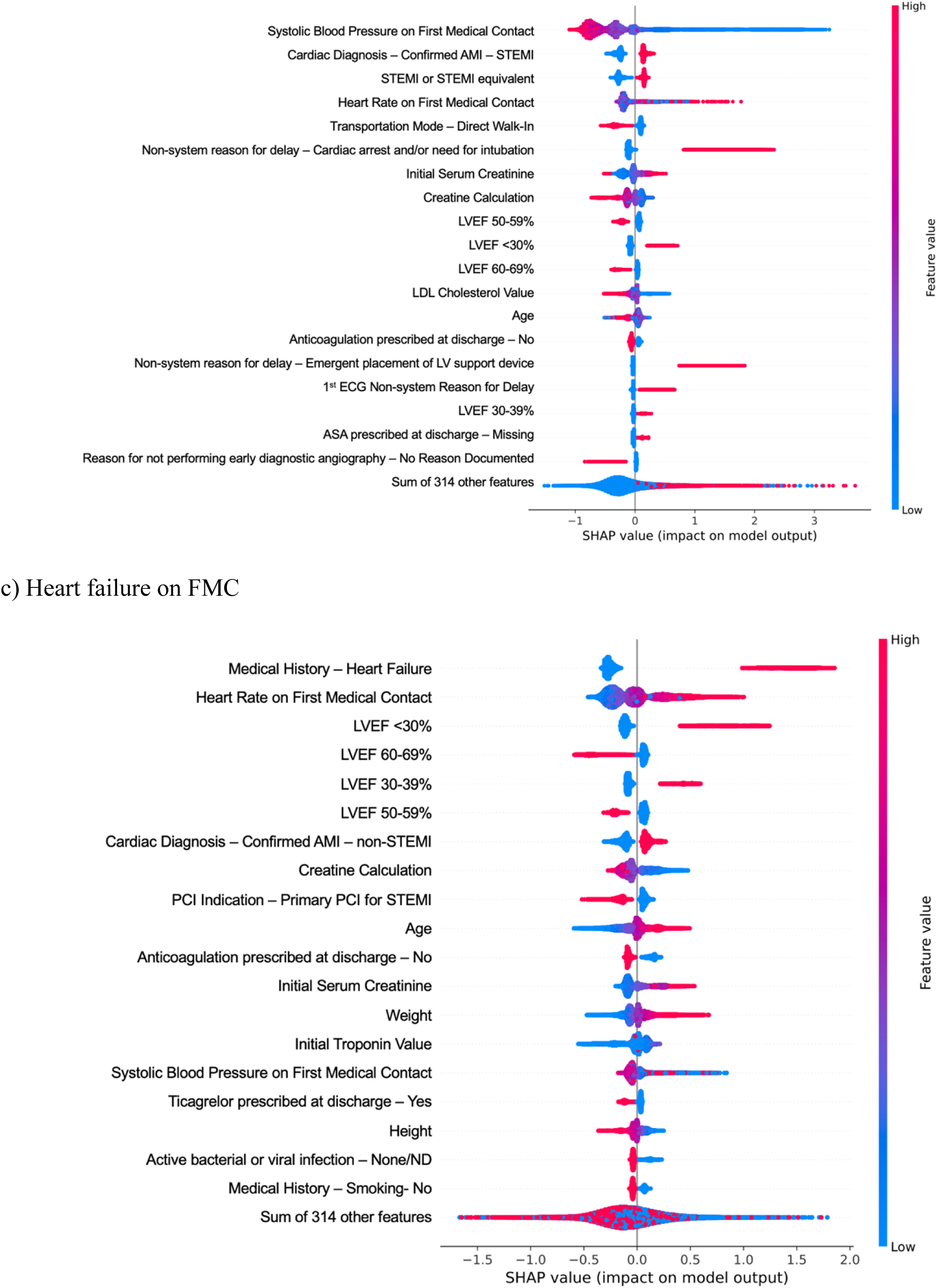
Top 20 features contributing for predicting outcomes by XGBoost model according to SHAP. In this set of beeswarm plots, each dot corresponds to an individual person from the training set. The dot’s position on the x-axis shows the impact that feature has on the model’s prediction for that person. For continuous variables, higher value is represented by red, while lower values by blue. For binary variables “yes” is indicated by red and “no” by blue. Where multiple dots land at the same x position, they stack to show density. Figure 4a depicts beeswarm plot for cardiac arrest prior to arrival; Figure 4b for cardiogenic shock on FMC; and Figure 4c for heart failure on FMC. *PCI* percutaneous coronary intervention; *FMC* first medical contact; *MI* myocardial infarction; *AMI* acute myocardial infarction; *STEMI* ST elevation myocardial infarction; *LVEF* left ventricular ejection fraction.

For predicting cardiac arrest prior to arrival, several features such as extreme heart rate measured on first medical contact, non-system reason for delay due to cardiac arrest and/or need for intubation, and cardiac diagnosis not stated as confirmed AMI - non-STEMI were all strong predictors (Figure 3a). Additionally, high serum creatinine levels, low systolic blood pressure on first medical contact, and younger age also positively contributed to the model prediction. The initial troponin value, non-system reasons for delay in the first ECG, presence of STEMI or STEMI equivalent, creatine calculation, and diabetes mellitus were also important predictors.

For predicting the cardiogenic shock on FMC (Figure 3b), we identified low systolic blood pressure on first medical contact as the most important predictor. Cardiac diagnosis of Confirmed AMI – STEMI, and STEMI or STEMI equivalent were strong predictors of the model. Other significant predictors include heart rate on first medical contact, mode of arrival (walk-in), non-system reason for delay due to cardiac arrest and/or in need for intubation, and lab values such as initial serum creatinine, creatinine calculation and Left Ventricular Ejection Fraction (LVEF) percentage.

For predicting heart failure on FMC (Figure 3c), medical history of heart failure, higher heart rate on FMC, and Left Ventricular Ejection Fraction (LVEF) percentage were the top three predictors. Specifically, LVEF below 40% increased the likelihood of presenting heart failure on FMC, whereas LVEF above 50% was indicative of absence. Other important predictors included age, initial serum creatinine, creatinine calculation, cardiac diagnosis of Confirmed AMI – non-STEMI, PCI indication of primary PCI for STEMI, no anticoagulation prescribed at discharge, etc.

Utilizing the SHAP values, we detailed the influence of the top 10 features in two specific patient cases using XGBoost models for heart failure on FMC. In the first patient case (Figure S1a), the top predictive variables for heart failure on FMC are LVEF percentage, heart rate on FMC, and cardiac diagnosis of confirmed AMI-non - STEMI.

For the second instance (Figure S1b), the top variables that for predicting the patient without heart failure on FMC are LVEF percentage between 60-69%, heart rate on FMC, troponin value, and not having a medical history of heart failure. A detailed description of variables and their values can be found in supplementary figures.

### Subgroup analysis

The models showed sex-based differences in the predicted prevalence rates for cardiac conditions. Specifically, XGBoost model predicted a higher prevalence rate of cardiac arrest prior to arrival (Male: 5.6%, Female: 3.6%) in male compared to female individuals but yielded a lower prevalence rate of heart failure on FMC (Male: 15.5%, Female: 20.5%) in male compared to female individuals and cardiogenic shock on FMC (Male: 4.9%, Female:5.2%). For race/ethnicity subgroups, individuals who identify as non-Hispanic Black had the highest predicted prevalence rate of heart failure on FMC (21.4%). Detailed summary statistics can be found in Supplementary Table 4.

Our model also exhibits variability in performance across different sex and racial/ethnic groups. Specifically, the model has a higher false negative rate (FNR) in females compared to males for identifying cardiac arrest prior to arrival (0.48 vs. 0.42), cardiogenic shock (FNR, 0.50 vs. 0.43), and heart failure (0.46 vs. 0.45), indicating a greater likelihood of missed true cases. Females also have a higher false positive rate (FPR) in cardiogenic shock (0.283 vs. 0.276) and heart failure (0.15. vs. 0.10). Among racial/ethnic groups, for cardiogenic shock, individuals who identify as Non-Hispanic Black were most affected, with the highest FNR of 0.53. Non-Hispanic Black patients also has the highest FNR (0.58) in cardiac arrest and the highest false positive rate (FPR) (0.14) in heart failure comparing to other groups. In addition, individuals who identify as Hispanic had the highest FPR (0.02) in cardiac arrest, suggesting a higher probability of incorrectly identifying conditions that were not present.

## DISCUSSION

Missing patients’ medical conditions commonly occurs in clinical dataset, which has resulted in discrepancies and limited the data power and quality.^20,21^ This study utilized multiple machine learning models to predict the presence of cardiac arrest, cardiogenic shock, and heart failure in patients arriving at hospitals with AMI. The advanced machine learning models including XGBoost and LightGBM demonstrated high predictive accuracy for identifying these three complications. Furthermore, by applying a novel framework SHAP, the study provides a transparent process to explain how the models predict the clinical conditions in a population scale and at the individual patient level. Although we used the national GWTG registry and the three conditions as the use case, the methodology and approach were not limited to those registries and conditions and can be generalized to phenotypes or clinical history that might be miss captured in any clinical dataset.

Previous studies have shown that XGBoost and LightGBM models can achieve strong imputation performance on commonly measured continuous laboratory variables such as chloride, potassium, creatinine, and glucose by leveraging both longitudinal and cross-sectional context in clinical datasets. ^4,9,22^ Our study adds to the existing literature by demonstrating that both models were also able to achieve strong performance for imputing binary (yes-no) clinical variables describing presenting patient characteristics such as cardiac arrest prior to arrival, cardiogenic shock on FMC and heart failure on FMC.

The improvement in imputation performance could be attributed to the diverse variety of patient characteristics and procedures during hospitalization in the GWTG data, enabling the models to infer missing values of clinical indicators from other non-missing variables collected in the registry. For instance, SHAP analysis shows that models for imputing cardiac arrest prior to arrival could infer information from non-system reasons for delay due to cardiac arrest and/or in need for intubation. Patients who have this particular reason for delay are much more likely to have cardiac arrest prior to arrival. Models for imputing cardiogenic shock on FMC also utilized the transportation mode information of the patients, with “direct walk in” patients being very unlikely to have cardiogenic shock on FMC. By extracting these features with complementary information from the registry, ML-based imputation models can achieve higher performance scores overall.

Some features influence imputation model outputs in unexpected ways based on the results of SHAP analysis. One of the most notable counterintuitive examples is that age is negatively associated with the patient having cardiac arrest prior to arrival. Further examination reveals that the median age of patients in the dataset who had cardiac arrest prior to arrival (63) is lower than the median age for those who do not (67), and that age is negatively correlated with cardiac arrest prior to arrival, with statistical significance (ρ = -0.085, p < 0.001). One possible explanation is that the registry is skewed towards older patients, with the relatively younger patients collected having more serious conditions.

Regarding model performance in different sex and race/ethnicity subgroups, we observed that female groups and non-Hispanic black groups are more likely to have missed true cases (i.e. presented with clinical indicators but predicted absence), while Hispanic groups are more likely to have been incorrectly identified as having true cases (i.e. absence in clinical indicators but predicted presence). This finding was consistent with a previous study that utilized ML models for predicting clinical outcomes in that female patients and underrepresented minority patients are more likely to experience under-prediction of clinical outcomes. ^23^

### Limitations

This study also had limitations. First, our ML-based imputation models used a large number of features, which may not be fully captured in some patients’ records. Imputation on missing features needs to be done prior to applying the model. Future research could focus on limiting the features used in these ML based models to most influential ones identified by SHAP while preserving model performance. Second, we did not include the clustering effects of hospitals and we dropped hospital level characteristics during the data processing stage for simplicity. If hospital-level characteristics influenced clinical indicators like cardiac arrest prior to arrival, then these effects may be unaccounted for and should be addressed in the future by incorporating these characteristics in analyses. s

## Conclusions

This study demonstrates that machine learning models, especially the XGBoot and LightGBM can accurately impute missing key clinical predictors – cardiac arrest, cardiogenic shock, and heart failure in patients with AMI. The novel use of SHAP in our analysis contributes to a transparent and explainable machine learning process which can provide further insights on the data management. Our findings also highlight certain variability in imputation accuracy across different sex and race/ethnicity groups, suggesting a need for future investigation. Despite focusing on a national registry and specific conditions, our approach is generalizable and could improve missing data imputation across various clinical datasets and conditions, thus enhancing the quality of clinical dataset.

## Data Availability

Data cannot be share publicly because of data use agreements of the American Heart Association. Deidentified data from the AHA GWTG-CAD regisrtry are available through the AHA Precision Medicine Platform. Requests to access the data should be made by contacting AHA as instructed at https://www.heart.org/en/professional/quality-improvement/quality-research-and-publications/hospital-level-research.

